# Croup during the COVID-19 Omicron Variant Surge

**DOI:** 10.1101/2022.02.02.22270222

**Authors:** Emine M. Tunҫ, Cassandra Koid Jia Shin, Etiowo Usoro, Siobhan E. Thomas-Smith, Indi Trehan, Russell T. Migita, Ashley E. Keilman

**Author notes:** **Address correspondence to:** Ashley Keilman, MD, Division of Pediatric Emergency Medicine, 4800 Sand Point Way NE, M/S MB.7.520, Seattle, WA 98105. **Contributors’ Statement Page** Drs. Tunç, Koid Jia Shin, and Siobhan-Thomas conceptualized and designed the study, drafted the manuscript, and revised the manuscript for important intellectual content. Mr. Usoro and Dr. Trehan conceptualized and designed the study, performed data acquisition, analysis, and interpretation, drafted the manuscript, revised the manuscript for important intellectual content, and take responsibility for the integrity of the data and the accuracy of the data analysis. Drs. Migita and Keilman provided administrative support, conceptualized and designed the study, drafted the manuscript, and revised the manuscript for important intellectual content. All authors approved the final manuscript as submitted.

## Abstract

Croup is a common upper respiratory disease usually associated with parainfluenza virus, resulting in stridor, hoarse voice, barky cough, and variable respiratory distress. Here we examine the data at our center confirming a sharp increase in cases of croup associated with the Omicron variant. Data was retrospectively extracted from patient charts among those seen in the Emergency Department at Seattle Children’s Hospital. Inclusion criteria were patients who were assigned a diagnosis containing “croup” during either 5/30/2021-11/30/2021, a time period correlating with predominance of the COVID-19 Delta variant (B.1.617.2), or the initial phase of the Omicron variant surge (12/1/2021-1/15/2022). Contemporaneous publicly available local data on the proportion of SARS-CoV-2 samples in surrounding King County, Washington, with spike gene target failure on TaqPath PCR assays was used as a proxy for the proportion of infections caused by the Omicron variant. A total of 401 patients were diagnosed with croup during the Delta surge and 107 patients were diagnosed with croup during the Omicron surge. Patients who presented during the Omicron surge were more likely to test positive for COVID-19 (48.2% vs 2.8%, p < 0.0001). Children with a clinical diagnosis of croup during the Omicron surge were more likely to be prescribed racemic epinephrine as part of their care (21.5% vs 13.0%, p = 0.032). There were no differences in presenting age, rate of admission, rate of return to the ED within 72 hours, or admission among those who returned within 72 hours. During the Omicron surge, the incidence of croup nearly doubled compared to the rate in prior months, while at the same time the number of cases of parainfluenza virus identified decreased. Consistent with prior case reports, we have identified a sharp rise in cases of croup seen in our pediatric ED in parallel with the replacement of the SARS-CoV-2 Delta variant by Omicron as the dominant variant in our community.

## Introduction

Croup (viral laryngotracheitis) is a common childhood upper respiratory disease, usually associated with parainfluenza or other endemic viruses, including seasonal coronavirus.^1^ The infection results in varying degrees of inspiratory stridor, hoarse voice, barky cough, and respiratory distress up to and including complete airway obstruction. Sporadic case reports have suggested an association between SARS-CoV-2 and croup.^2–9^ *Ex vivo* studies suggest that the Omicron variant (B.1.1.529) of SARS-CoV-2 replicates more rapidly in higher airways than previous lineages, suggesting the possibility of an increased risk for croup.^10^ Here we merge epidemiologic and clinical data to evaluate whether there is indeed a high rate of croup as a manifestation of COVID-19, specifically with the Omicron variant.

## Methods

High-level data was retrospectively extracted from patient charts among those seen in the Emergency Department (ED) at Seattle Children’s Hospital, an academic, quaternary-care hospital with more than 50,000 annual ED visits prior to the COVID-19 pandemic. Inclusion criteria were patients who were assigned a diagnosis containing the word “croup” during either 5/30/2021-11/30/2021, a time period correlating with predominance of the SARS-CoV-2 Delta variant (B.1.617.2), or during 12/1/2021-1/15/2022, corresponding to the initial phase of the Omicron variant surge.

Qualitative SARS-CoV-2 polymerase chain reaction (PCR) testing from nasopharyngeal or mid-turbinate swabs were obtained at the discretion of the treating clinician, as were interventions including racemic epinephrine and hospitalization. Contemporaneous publicly available local data on the proportion of SARS-CoV-2 samples in surrounding King County, Washington, with spike gene target failure on TaqPath PCR assays was used as a proxy for the proportion of infections caused by the Delta and Omicron SARS-CoV-2 variants (https://depts.washington.edu/labmed/covid19/#sequencing-information). This study was approved by the Seattle Children’s Hospital Institutional Review Board.

## Results

A total of 401 patients were diagnosed with croup during the Delta surge and 107 patients were diagnosed with croup during the Omicron surge. Patients with croup who presented during the Omicron surge were more likely to test positive for COVID-19 (48.2% vs 2.8%, p < 0.0001, odds ratio 31.8) (**Table 1**). The rate of viral respiratory testing remained similar during both time periods. Children with a clinical diagnosis of croup during the Omicron surge were more likely to be administered racemic epinephrine as part of their care (21.5% vs 13.0%, p = 0.032). There were no differences in presenting age, rate of admission, rate of return to the ED within 72 hours, or admission among those who returned within 72 hours.

**Table 1.** Characteristics of croup patients during two different phases of the COVID-19 pandemic.

| **Characteristic** | **COVID Phase** |  | **p-value<sup>c</sup>** | **Odds ratio (95% CI)** |
| --- | --- | --- | --- | --- |
|  | **Delta Surge<sup>a</sup>**<br>N = 401 | **Omicron Surge<sup>b</sup>**<br>N = 107 |  |  |
| Age, median (IQR), yr | 2.33 (1.42, 3.83) | 2.25 (1.08, 3.62) | 0.29 |  |
| SARS-CoV-2 status, n (%) |  |  |  |  |
| Detected | 6 (2.8%) | 27 (48.2%) | < 0.0001 | 31.8 (12.7-80.2) |
| Not detected | 205 (97.2%) | 29 (51.8%) |  |  |
| Not tested | 190 | 51 |  |  |
| Received racemic epinephrine, n (%) | 52 (13.0%) | 23 (21.5%) | 0.032 | 1.84 (1.06-3.18) |
| Admitted to hospital, n (%) | 15 (3.7%) | 4 (3.7%) | > 0.99 | 1.00 (0.35-2.85) |
| 72-hr return to ED, n (%) | 12 (3.0%) | 6 (5.6%) | 0.23 | 1.93 (0.72-5.06) |
| 72-hr return admission, n (%) | 3 (25.0%) | 1 (16.7%) | > 0.99 | 0.60 (0.04-5.19) |

<sup>a</sup> Time period when Delta variant was locally predominant: 5/30/2021 – 11/30/2021.
<sup>b</sup> Time period when Omicron variant was locally predominant: 12/1/2021 – 1/15/2022.
<sup>c</sup> Wilcoxon rank sum test for age; Fisher's exact test for other variables.

Using the local community prevalence of spike gene target failure as a proxy for the Omicron variant, the incidence of croup double compared to the rate in prior months, while at the same time the number of cases of parainfluenza virus identified decreased (**Figure 1**).

**Figure 1.**
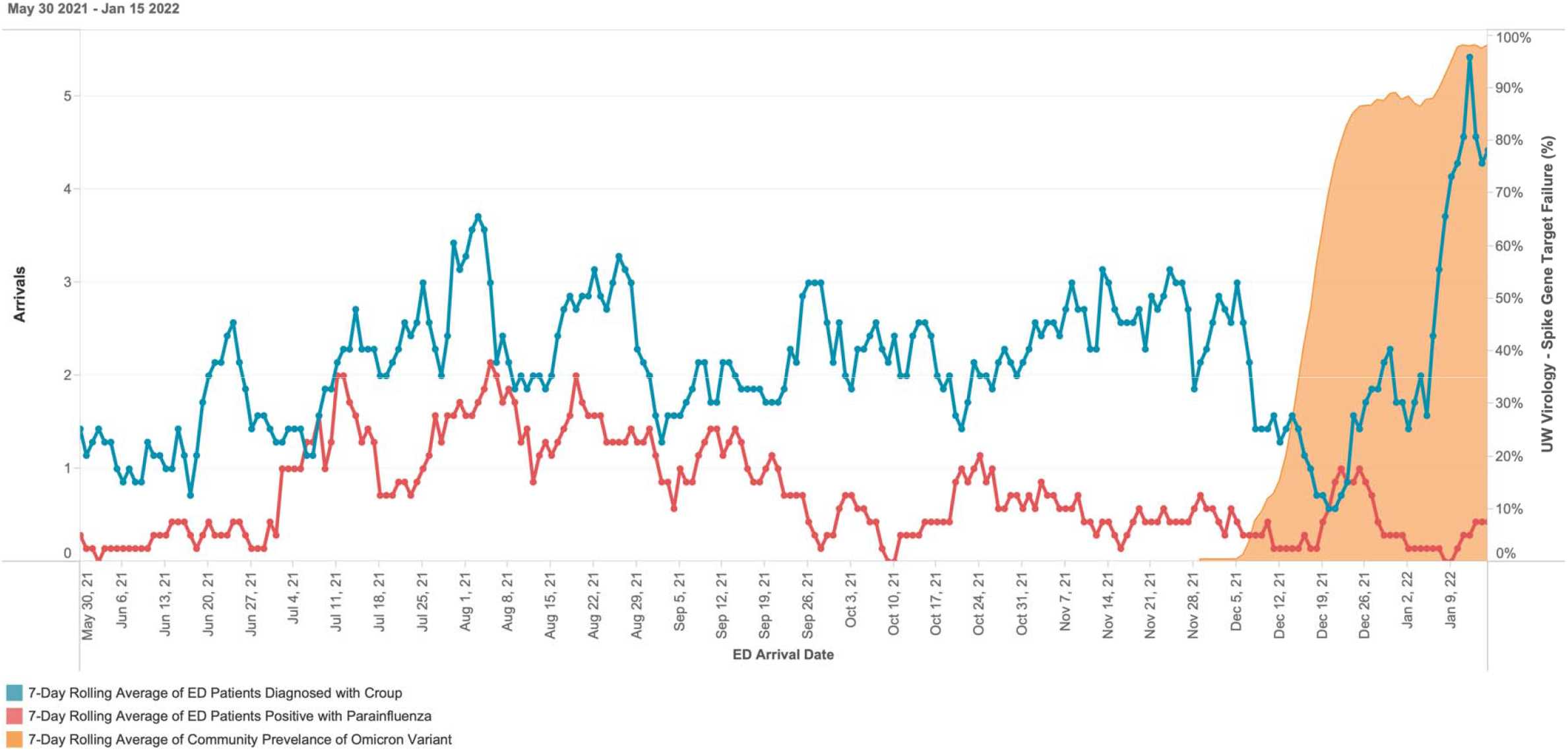
Number of patients diagnosed with croup, number of patients diagnosed with parainfluenza, and community prevalence of spike gene target failure for SARS-CoV-2 (7-day rolling averages).

## Discussion

We have identified a sharp rise in cases of croup seen in our pediatric ED in parallel with the replacement of the SARS-CoV-2 Delta variant by Omicron as the dominant variant in our community. This increase was not temporally associated with the circulation of parainfluenza virus, and indeed Omicron replaced parainfluenza as the predominant virus associated with croup during this time. Viral testing is not routinely recommended for patients with croup; testing that was performed was for a wide variety of patients and indications, but we do believe our data is a reasonable proxy for local population prevalence. The end of the study period coincided with a decrease in hospital testing capacity and thus we were not able to see if this trend continued with as much precision.

Croup patients during the Omicron surge were more likely to receive racemic epinephrine, suggesting a more severe initial clinical presentation. Based on admission rates and return visits, there is not enough data to alter current management after ED discharge.

Croup is one of the most severe manifestations of upper respiratory infections in children. As a common reason for ED visits, croup places a significant resource burden on health care systems. It appropriately generates tremendous fear and anxiety among children and their caretakers. As has been seen repeatedly during the COVID-19 pandemic, the protean clinical manifestations of SARS-CoV-2 have frequently humbled health care providers, with croup now needing to be recognized as a common presentation of the Omicron variant, and possibly future variants as well.

## Data Availability

All data produced in the present work are contained in the manuscript.

## Abbreviations

COVID-19: coronavirus disease 2019
ED: Emergency Department
PCR: polymerase chain reaction
SARS-CoV-2: severe acute respiratory syndrome coronavirus 2

## Acknowledgments

We thank Pavitra Roychoudhury, PhD, Department of Laboratory Medicine and Pathology, University of Washington, for her help in accessing local SARS-CoV-2 surveillance data.

## Notes

**Conflict of interest disclosures:** None.

**Funding support:** None.

### Competing Interest Statement

The authors have declared no competing interest.

### Funding Statement

The study did not receive any funding.

### Author Declarations

IRB of Seattle Children’s Hospital waived ethical approval for this work.

## References

1. Sung JY, Lee HJ, Eun BW, Kim SH, Lee SY, Lee JY, et al. Role of human coronavirus NL63 in hospitalized children with croup. Pediatr Infect Dis J. 2010;29:822–826.

2. Brackel CLH, Rutjes NW, Kuijpers TW, Terheggen-Lagro Swj. SARS-CoV-2 and croup, a rare relationship or coincidence? Am J Emerg Med. 2021;49:410–411.

3. Kaur R, Schulz S, Fuji N, Pichichero M. COVID-19 pandemic impact on respiratory infectious diseases in primary care practice in children. Front Pediatr. 2021;9:722483.

4. Lim CC, Saniasiaya J, Kulasegarah J. Croup and COVID-19 in a child: A case report and literature review. BMJ Case Rep. 2021;14.

5. Peterson K, Patel J, Collier C, Chan SB. SARS-CoV-2 and croup, not a rare coincidence. Am J Emerg Med. 2021;doi:10.1016/j.ajem.2021.12.023.

6. Pitstick CE, Rodriguez KM, Smith AC, Herman HK, Hays JF, Nash CB. A curious case of croup: Laryngotracheitis caused by COVID-19. Pediatrics. 2021;147:e2020012179.

7. Tsoi K, Chan KC, Chan L, Mok G, Li AM, Lam HS. A child with SARS-CoV2-induced croup. Pediatr Pulmonol. 2021;56:2377–2378.

8. Venn AMR, Schmidt JM, Mullan PC. Pediatric croup with COVID-19. Am J Emerg Med. 2021;43:287.e1-287.e3.

9. Zuccarelli AM, Leonard CG, Hampton SM. Laryngotracheobronchitis, croup, an unusual presentation of SARS-CoV-2. Ulster Med J. 2022;91:57–58.

10. Chan MCW, Hui KPY, Ho J, Cheung M, Ng K, Ching R, et al. SARS-CoV-2 omicron variant replication in human respiratory tract ex vivo. Nature Portfolio. 2021;doi:10.21203/rs.3.rs-1189219/v1.

